# Leveraging Machine Learning Models and Pharmacy Refill Adherence as a Cost-Effective Proxy for Predicting HIV Viral Suppression during Antiretroviral Therapy in Resource-Limited Settings

**DOI:** 10.64898/2026.01.05.26343496

**Authors:** Meshack D. Lugoba, Raphael Z. Sangeda, Ritah F. Mutagonda, James Mwakyomo, George Musiba, Veryeh Sambu, Beatrice Mutayoba, Mercy Mpatwa, Prosper Njau, Werner Maokola

## Abstract

**Introduction:** Achieving viral suppression is central to HIV epidemic control; however, routine viral load (VL) testing in many low- and middle-income countries is constrained by laboratory capacity, logistics, and cost. In Tanzania, disparities in VL coverage persist across age groups and geographical regions, limiting the timely detection of treatment failure. Pharmacy refill adherence is a low-cost, routinely collected objective indicator of treatment behavior. This study assessed whether pharmacy refill adherence, enhanced using machine learning (ML) models, can reliably predict viral suppression among people living with HIV (PLHIV) in Tanzania.

**Methods:** We conducted a retrospective analysis using nationally representative patient-level data from the Care and Treatment Center (CTC-2) database, collected between 2017 and 2021. A random sample of 40,000 records was drawn, of which 28,044 patients met the inclusion criteria. Pharmacy refill adherence was calculated as the proportion of days covered and capped at 100%. Viral suppression was defined as a VL of <1,000 copies/mL. Logistic regression, Random Forest, Gradient Boosting Machine (GBM), and XGBoost models were trained using an 80/20 training–testing split, and the model performance was evaluated using the area under the receiver operating characteristic curve (AUC). Youden’s Index was used to determine the optimal adherence threshold.

**Results:** Among the 28,044 patients included in the analysis, the median age at ART initiation was 38 years, and 64.9% were female. The median pharmacy refill adherence was 90.64% (mean, 87.37%). Viral load (VL) measurements were available for 21,572 patients, of whom 88.7% achieved viral suppression. Higher pharmacy refill adherence was strongly associated with viral suppression, whereas lower adherence was observed among adolescents, young adults, and individuals who were lost to follow-up. Marked geographic variation was observed, with higher adherence in regions such as Dar es Salaam and lower adherence in more remote regions, including Rukwa and Singida. Among machine learning models, XGBoost demonstrated the highest predictive performance (AUC >0.85), followed by Gradient Boosting Machines and Random Forest, while logistic regression provided stable baseline estimates. Pharmacy refill adherence, duration of follow-up, clinic visit frequency, and patient age were the strongest predictors of viral suppression.

**Conclusion:** Pharmacy refill adherence is a strong predictor of viral suppression and provides a feasible and cost-effective tool for monitoring ART outcomes in settings with limited VL testing. Machine learning approaches further enhance the predictive value of routine program data and can support the early identification of patients at risk of virological failure. Integrating adherence-based predictive analytics into national HIV program monitoring systems may strengthen differentiated service delivery, improve treatment outcomes, and accelerate progress toward the UNAIDS 95–95–95 targets in Tanzania and similar resource-limited settings.

## Introduction

Human immunodeficiency virus (HIV) infection remains a major global public health challenge worldwide. In 2024, an estimated 40.8 million people were living with HIV worldwide, and approximately 0.6 million deaths were attributed to HIV-related causes (World Health Organization, 2025). Achieving the Sustainable Development Goal of ending AIDS as a public health threat by 2030 requires strengthened HIV prevention, expanded access to antiretroviral therapy (ART), and sustained viral suppression among people living with HIV (UNAIDS, 2016; World Health Organization, 2025).

The global expansion of ART has contributed to substantial declines in HIV incidence and mortality over the past decade. To accelerate epidemic control, the Joint United Nations Programme on HIV/AIDS (UNAIDS) introduced the 95–95–95 targets in 2014, aiming for 95% of people living with HIV to know their status, 95% of those diagnosed to receive sustained ART, and 95% of those on ART to achieve viral suppression (Keiser et al., 2011; UNAIDS, 2016). Achieving the third "95" depends on reliable viral load (VL) monitoring, which the World Health Organization (WHO) recommends as the gold standard for assessing treatment response and identifying virological failure (World Health Organization, 2025). Compared with immunological or clinical monitoring, VL testing is more sensitive and specific, enabling earlier detection of treatment failure, reducing mortality, improving outcomes, and lowering the risk of HIV drug resistance (Estill et al., 2018; Piot & Quinn, 2013; Sollis et al., 2014).

Despite its importance, VL monitoring remains limited in many low- and middle-income countries (LMICs) due to financial constraints, limited laboratory capacity, inconsistent sample transport systems, and shortages of trained personnel (Lecher et al., 2021; Roberts et al., 2016; Rutstein et al., 2016). These challenges contribute to suboptimal VL coverage, delayed clinical action following elevated VL results, and a persistent risk of undetected treatment failure. In Tanzania, although the Ministry of Health adopted WHO’s "test-and-treat" guidelines in 2019 (United Republic of Tanzania, 2019), disparities in access to VL testing persist, particularly among adolescents, young adults, women, and patients attending remote Care and Treatment Centers (CTCs) (Amour et al., 2022; Martelli et al., 2019).

Given these limitations, there is a growing need for alternative or complementary approaches to identify patients at risk of virological failure, especially in settings where VL testing is infrequent or delayed. Pharmacy refill adherence is a promising, low-cost metric with a demonstrated predictive value for HIV treatment outcomes. Evidence from Tanzania and other LMICs indicates that pharmacy refill tracking outperforms self-reported adherence measures in predicting virological suppression and can serve as an early warning indicator of treatment failure (Martin et al., 2017; Sangeda et al., 2014). Because refill data are routinely collected, objective, and inexpensive to analyze, they offer a practical option for monitoring adherence in resource-limited settings.

However, adherence behavior is complex and shaped by patient, caregiver, family, societal and health-system-level factors (Haberer & Mellins, 2009; Nichols et al., 2017). Children, adolescents, and young adults face additional challenges, including poverty, food insecurity, caregiver dependence, stigma, and barriers to accessing health services, which often result in suboptimal adherence, lower rates of viral suppression, and an increased risk of HIV drug resistance(Lowenthal et al., 2014; Muri et al., 2017; Tabb et al., 2018). These vulnerabilities underscore the need for improved tools to identify individuals at risk of treatment failure.

Recent studies suggest that machine learning (ML) approaches can further enhance the prediction of HIV treatment outcomes by leveraging routinely collected clinical and programmatic data. A multi-site analysis demonstrated that ML models could accurately predict viral suppression among people living with HIV, outperforming conventional statistical approaches (Yang et al., 2025). Similarly, a facility-based study from Ethiopia reported that ML algorithms effectively identified patients at high risk of virological failure using ART follow-up data (Mamo et al., 2023). Although prior research has established the utility of pharmacy refill metrics in Tanzania (Martelli et al., 2019; Sangeda et al., 2014), limited evidence exists on whether combining refill-based adherence metrics with ML models can improve the prediction of viral suppression on a national scale. Tanzania’s CTC-2 database, which contains nationwide longitudinal pharmacy, clinical, laboratory, and demographic data, provides a unique opportunity to address this question.

Therefore, this study investigated whether pharmacy refill adherence augmented with ML algorithms can serve as a cost-effective proxy for predicting viral suppression among people living with HIV in Tanzania. By identifying patients at the highest risk of virological failure, particularly in contexts with incomplete VL testing coverage, this approach may support differentiated service delivery, enable earlier intervention, and strengthen national progress toward achieving the UNAIDS 95–95–95 targets.

## Methodology

### Study Area

This study used data from Care and Treatment Centers (CTCs) located in 26 regions in Tanzania. These centers are integral to the country’s HIV care and treatment programs, which are coordinated by the National AIDS, Sexually Transmitted Diseases, and Hepatitis Control Program (NASHCOP).

### Study Design and Population

This retrospective study analyzed patient-level data from the NASHCOP CTC-2 database, which includes nationwide information on sociodemographic characteristics, clinical characteristics, ART regimens, laboratory tests, pharmacy refill records, and treatment outcomes. The study period was from January 1, 2017, to December 31, 2021, and included patients enrolled in ART treatment programs at CTCs. No age limit was applied to the inclusion criteria of this study.

### Inclusion and Exclusion Criteria

Inclusion criteria for the study were for patients enrolled in CTCs between the start date of 1^st^ July 2017 and the end date of 30th June 2021. The exclusion criteria were as follows: patients with only one visit date, without a calculated pharmacy refill adherence measurement, who never attended care 6 months post-start date, or who had a start visit date equal to the study end date.

### Sample Size

A simple random sample of 40,000 patients was drawn from a pool of approximately 1.5 million patients enrolled in CTCs during the study period. After excluding patients with missing or incomplete data, the final sample comprised 28,044 individuals. A total of 11,956 were removed due to a lack of calculated adherence measurement (3,149), only being visited once (2,787), being lost for 6 months before the study start date (1,095), or having their last visit after 30th June 2021 (1,832). The exclusion criteria may have overlapped.

### Data Collection

Data were collected from the NASHCOP CTC-2 database, which includes detailed records of each patient’s socio-demographics, ART regimen history, viral load measurements, pharmacy refill data, and treatment outcomes. The key variables used in the analysis included age, gender, marital status, region of residence, ART regimen type, adherence rates, and viral load results. Other factors, such as follow-up duration, number of therapy changes, and visit counts, were also included in the predictive models to assess their relationship with virological success.

Pharmacy refill adherence was the primary predictor variable, calculated as the percentage of prescribed ART doses collected by patients over the study period. Refill adherence was denoted as 100% if all pills were collected on time, whereas refill rates exceeding 100% (indicating early collection) were capped at 100%. The formula used to calculate refill adherence is as follows:

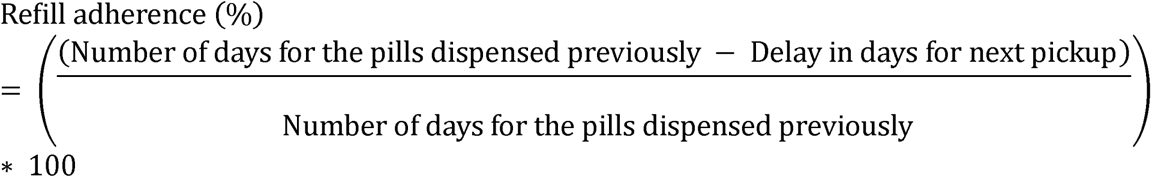

Patients were considered to have good adherence if their refill percentage was greater than or equal to the optimal cutoff determined by this analysis. In contrast, poor adherence was defined as a refill percentage below the optimal cutoff.

Viral load suppression (virological success) was defined as achieving a viral load of less than 1000 copies per milliliter, in accordance with the HIV and AIDS treatment guidelines in Tanzania. Virological failure was defined as the inability to achieve this level of viral suppression. All data were collected and managed using the R software for statistical analysis.

### Data Management and Statistical Analysis

The data were cleaned and preprocessed to exclude patients with incomplete records. Continuous variables were summarized using means and standard deviations, while categorical variables were summarized using frequencies and percentages.

Pharmacy refill adherence and selected clinical and programmatic variables, including follow-up duration, number of therapy changes, and visit counts, were analyzed using multiple predictive modeling approaches to assess their associations with virological outcomes. Several predictive models have been implemented for this purpose, including logistic regression, Random Forest, Gradient Boosting Machine (GBM), and Extreme Gradient Boosting (XGBoost). Logistic regression was used as a baseline model to assess the linear association between pharmacy refill adherence and virological outcomes. Random Forest models were applied to capture non-linear relationships and higher-order interactions using ensemble decision trees (Breiman, 2001).

GBM was employed to iteratively improve the prediction accuracy using sequential tree-based learning (Friedman, 2001). In contrast, XGBoost, an optimized and scalable boosting algorithm, was used to further model performance on large datasets (Chen & Guestrin, 2016).

The dataset was randomly split into a training set (80%) and a test set (20%). Model performance was evaluated using Receiver Operating Characteristic (ROC) curves, and predictive accuracy was quantified using the Area Under the ROC Curve (AUC). The optimal pharmacy refill adherence threshold for predicting virological success was determined using Youden’s Index, which maximizes the combined sensitivity and specificity.

To address the class imbalance in the outcome variable (88.7% of patients achieving virological success and 11.3% experiencing virological failure), class weighting was applied to the Random Forest and XGBoost models. Hyperparameter tuning was performed to optimize the model performance and improve the predictive stability.

### Ethical Considerations

Ethical clearance for this study was obtained from the Directorate of Research and Publication of the Muhimbili University of Health and Allied Sciences (MUHAS) with reference number DA.282/298/01.C/. The NASHCOP administration granted permission to collect the data. Strict privacy and confidentiality were maintained throughout the study. Only de-identified data were used, and patient IDs, names, or other personally identifiable information were not collected from the patients. Data collection adhered to national and international ethical standards, particularly regarding the handling of sensitive health information.

## Results

### Study Population

From the 40,000 sampled records, 28,044 patients met the inclusion criteria after excluding 3,149 without a calculable adherence measurement, 2,787 with a single clinic visit, 1,095 who did not return within six months of ART initiation, and 1,832 whose final visit occurred after the study end date. A total of 18,365 (65.5%) were female, while 9,679 (34.5%) were male (Table 1). The median age at ART initiation was 38 years (mean 37.82 years) (Table 2). Adults aged 29–69 years comprised 76.7% of the cohort (Table 1). The patients had a median follow-up duration of 1,473 days, with a median of 24 clinic visits and one therapy change (Table 2).

**Table 1:**
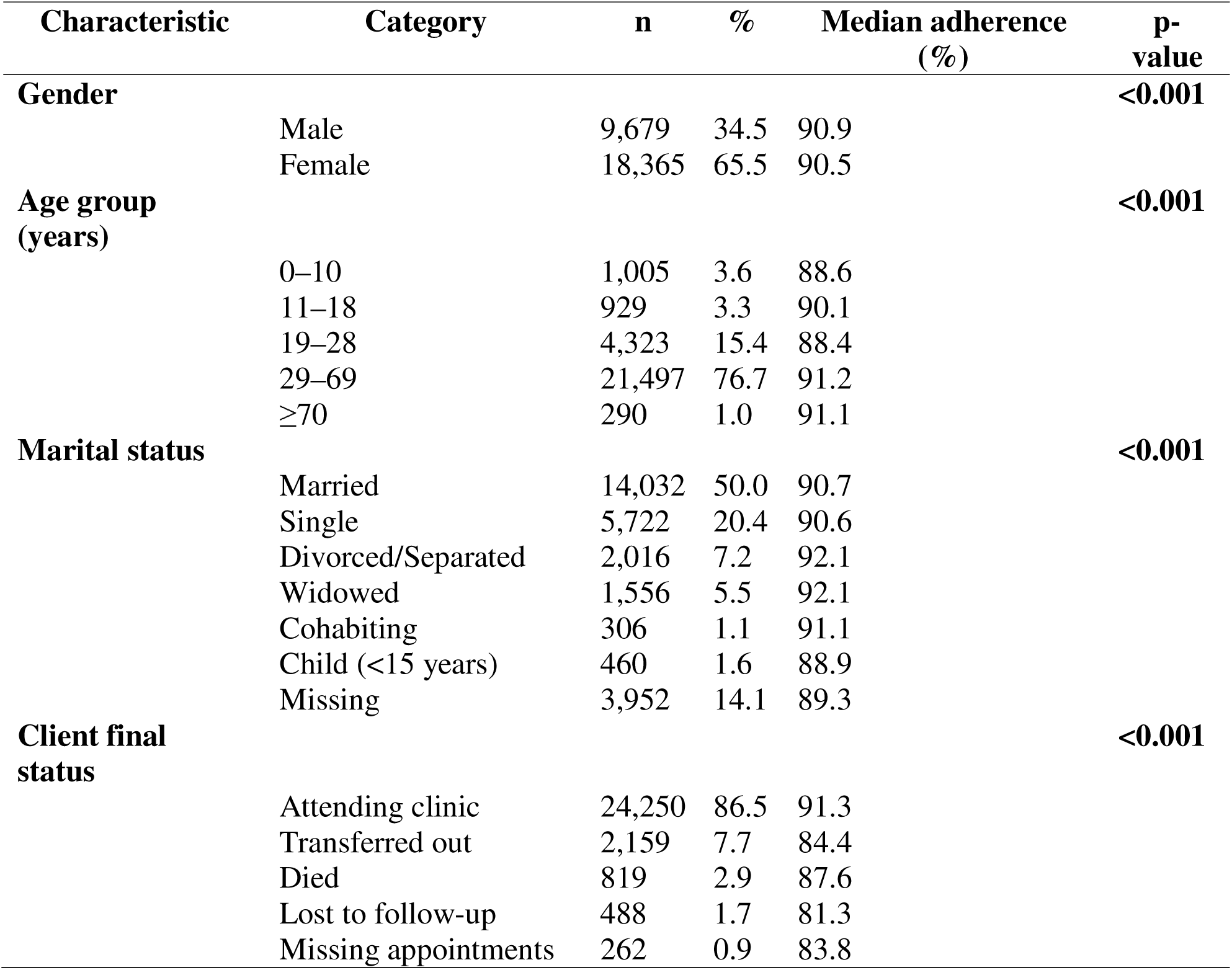
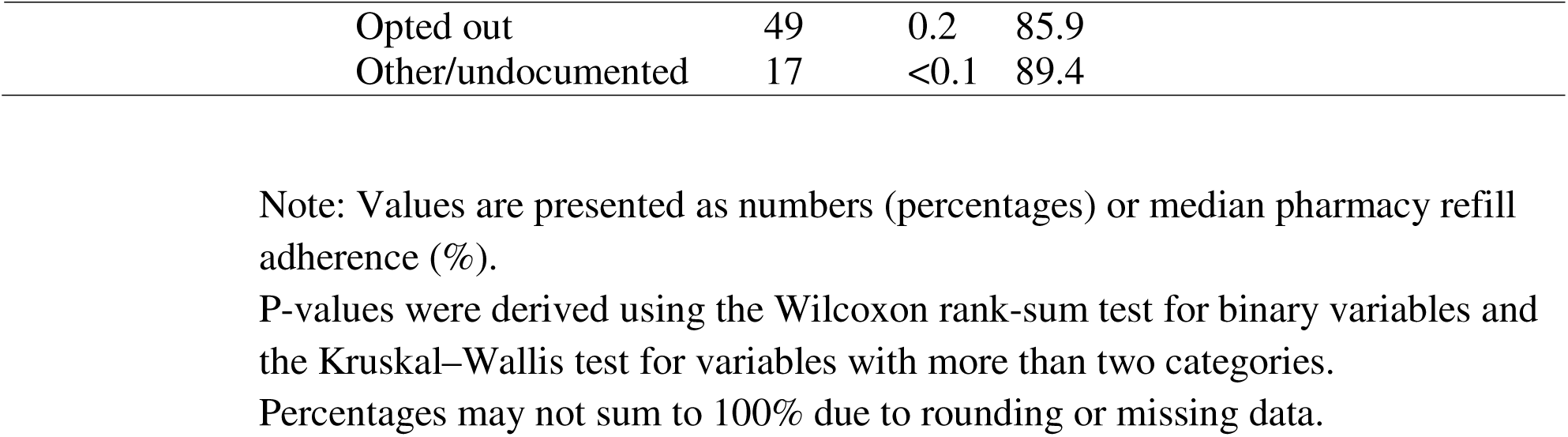
Baseline demographic and clinical characteristics and median pharmacy refill adherence among people living with HIV enrolled in Care and Treatment Centers in Tanzania, 2017–2021 (N = 28,044)

**Table 2:** Continuous baseline demographic and clinical characteristics of participants enrolled in HIV CTCs in Tanzania (2017–2021)

| Variable | Mean | Median | 95% CI<br>(Lower) | 95% CI<br>(Upper) | SD |
| --- | --- | --- | --- | --- | --- |
| Age at ART start (years) | 37.82 | 38 | 37.66 | 37.97 | 13.45 |
| Pharmacy refill adherence (%) | 87.37 | 90.64 | 87.23 | 87.52 | 12.58 |
| Number of clinic visits | 24.86 | 24 | 24.72 | 24.99 | 11.50 |
| Therapy change count | 1.14 | 1 | 1.13 | 1.15 | 0.87 |
| Follow-up duration (days) | 1254.48 | 1473 | 1248.19 | 1260.78 | 537.68 |
| Years under therapy | 3.44 | 4.04 | 3.42 | 3.45 | 1.47 |

In 2017, 4,346 (15.5%) patients were receiving dolutegravir (DTG)-based therapy at ART initiation, increasing to 25,458 (90.8%) by 2021 (Figure 1).

**Figure 1:**
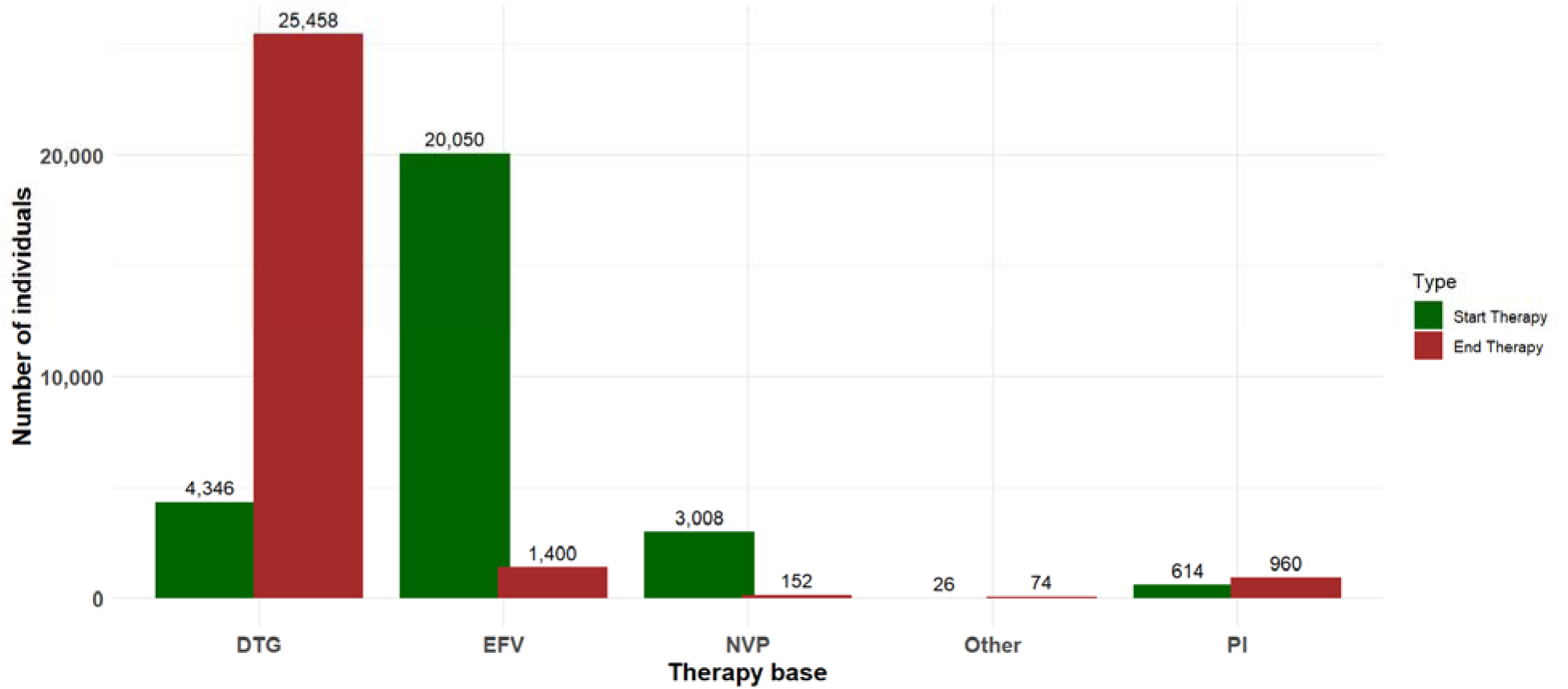
Change of therapy base at the start of the year 2017 and the end of the study period in 2021 DTG=Dolutegravir; EFV=efavirenz; NVP=Nevirapine, PI= Protease Inhibitor

### Pharmacy Refill Adherence

The median pharmacy refill adherence was 90.6% (mean 87.4%, SD 12.6). Adherence differed significantly by age group (p < 0.001), with lower levels observed among children aged 0–10 years (88.6%) and young adults aged 19–28 years (88.4%), and higher adherence among adults aged 29–69 years (91.2%) and older people aged ≥70 years (91.1%). Adolescents aged 11–18 years had a median adherence rate of 90.1% (Figure 2).

**Figure 2:**
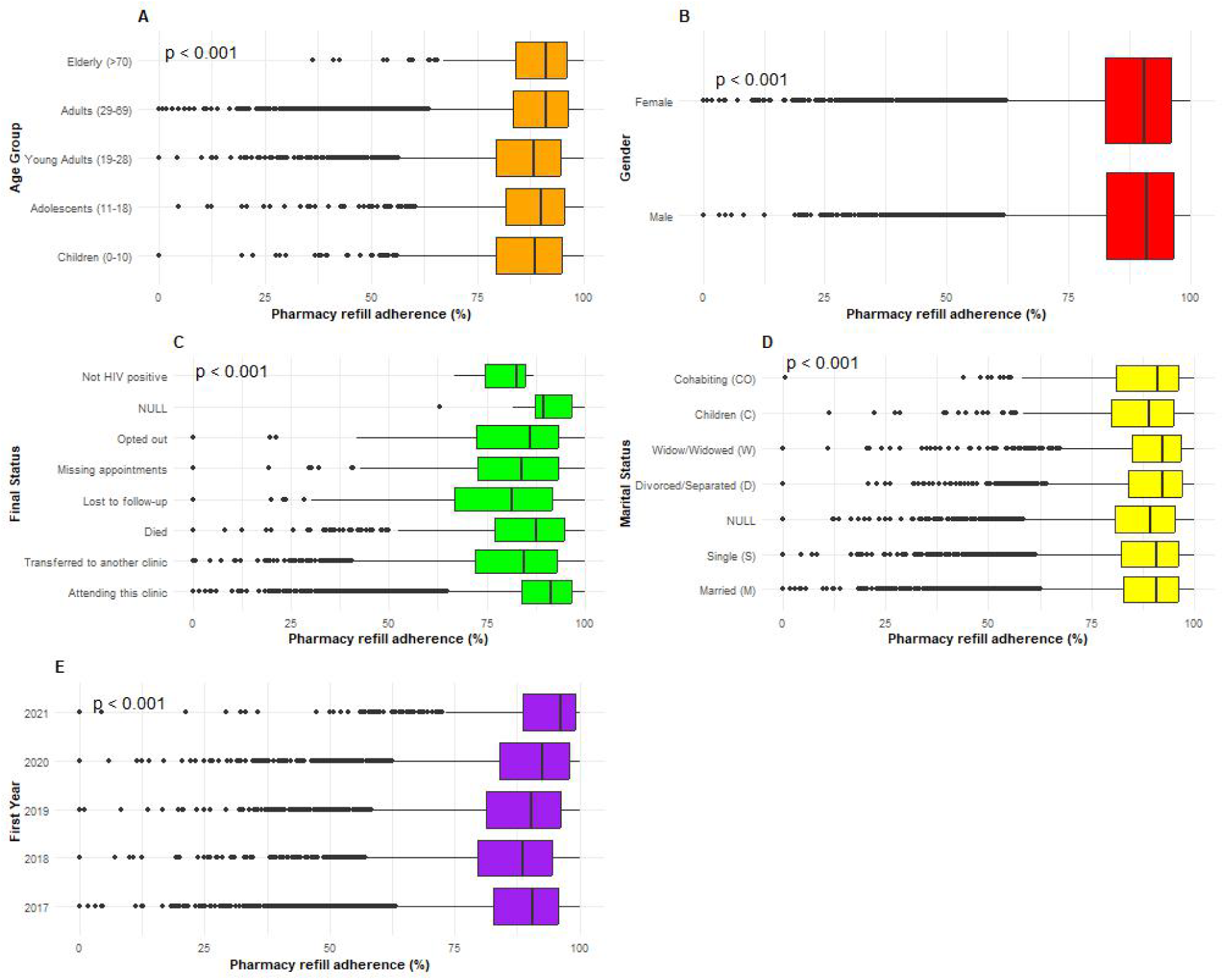
Pharmacy Refill Adherence (%) Across Demographic and Clinical Categories (Panels A: Age Group, B: Gender, C: Clinic Attendance, D: Marital Status and E: Year of Enrollment)

These age-related patterns are further illustrated by the continuous variation in adherence across single-year age increments, showing a clear transition from lower adherence among young adults aged 19–28 years to higher adherence among adults aged 29–69 years (Figure 3).

**Figure 3:**
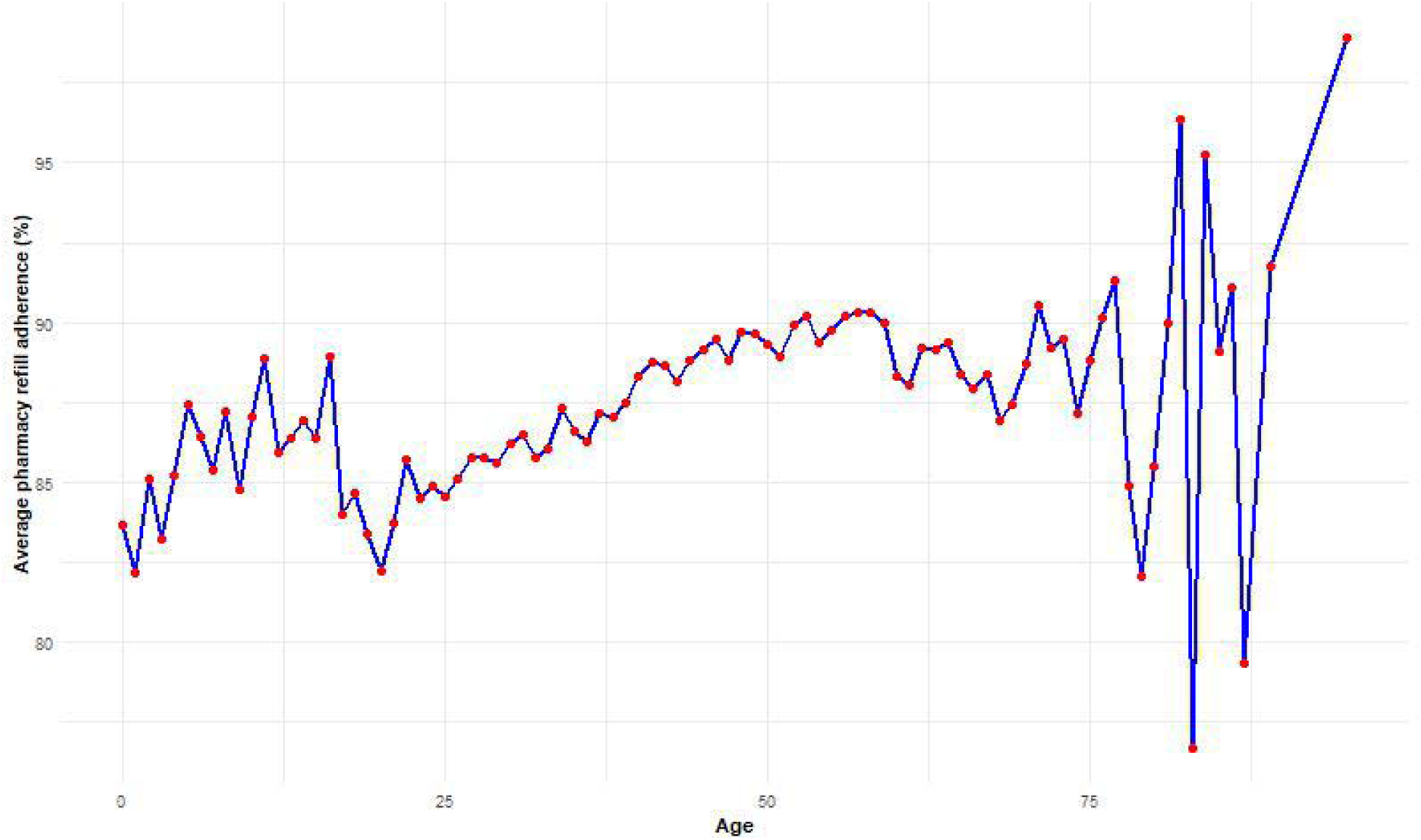
Pharmacy Refill Adherence (%) variation by single-year age increments among HIV individuals in Tanzania between 2017 and 2021

Males demonstrated slightly higher adherence (90.9%) than females (90.5%) (p < 0.001). Regarding clinical status, adherence was highest among patients actively attending the same clinic (91.3%) and lowest among those lost to follow-up (81.3%) (p < 0.001). By marital status, widowed and divorced patients had adherence levels exceeding 92%, while children (<15 years) had lower adherence (88.9%) (p < 0.001). Adherence also increased by calendar year of ART initiation, rising from 90.47% in 2017 to 96.3% in 2021 (p < 0.001) (Figure 2).

Regionally, median pharmacy refill adherence varied significantly across Tanzania (p < 0.001), ranging from 93.5% in Dar es Salaam and 93.4% in Iringa to 84.8% in Singida and 84.6% in Rukwa (Figure 4) and Supplementary Table 1. At the district level, adherence ranged from 95.35% in Karatu and 95.30% in Muheza to 78.5% in Liwale and 78.3% in Kalambo (P < 0.001). These patterns demonstrate substantial geographic heterogeneity in adherence across both regions and districts (Figure 5) and Supplementary Table 2).

**Figure 4:**
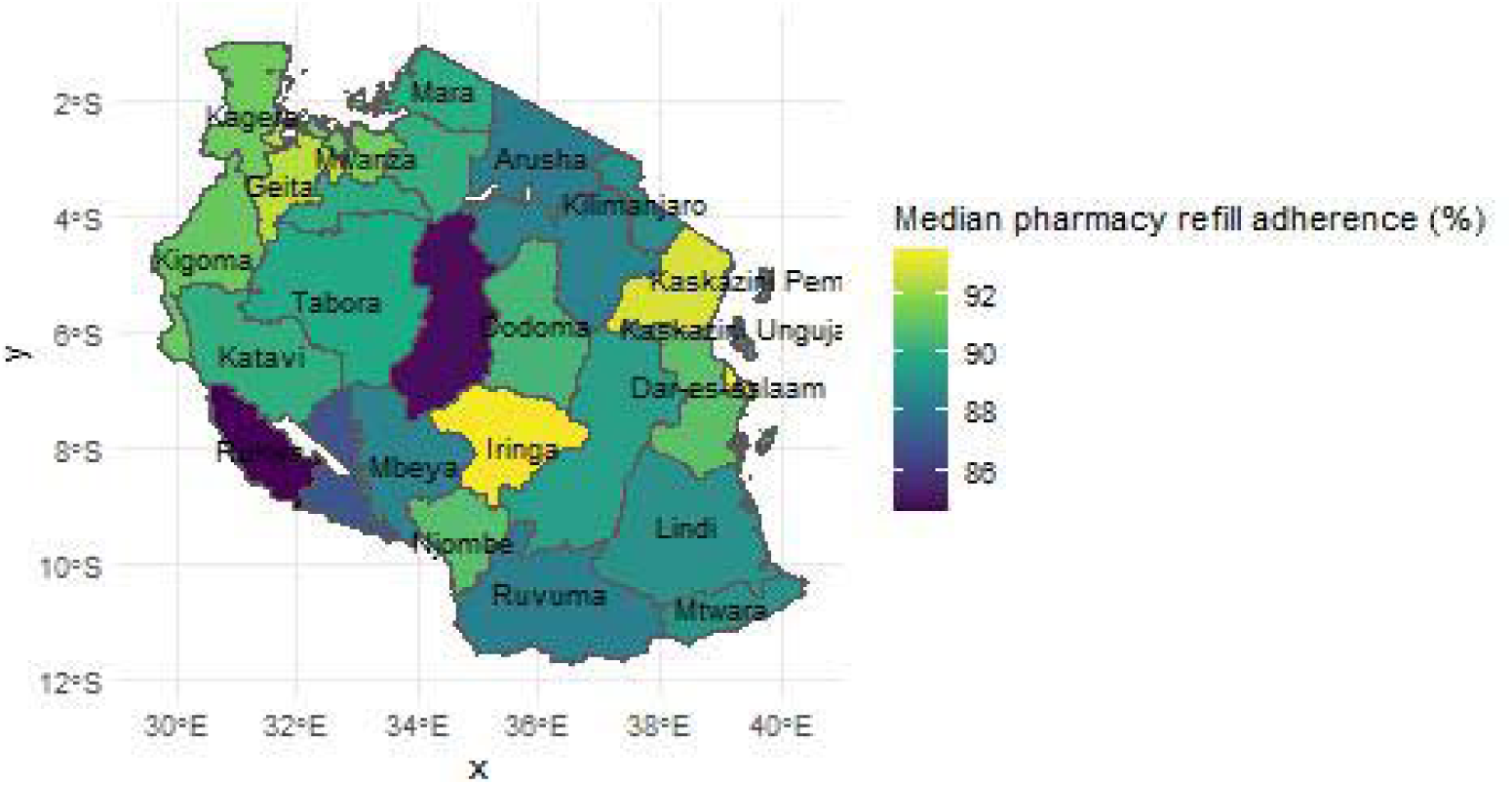
Geospatial Distribution of Pharmacy Refill Adherence (%) Across Regions of Tanzania

**Figure 5:**
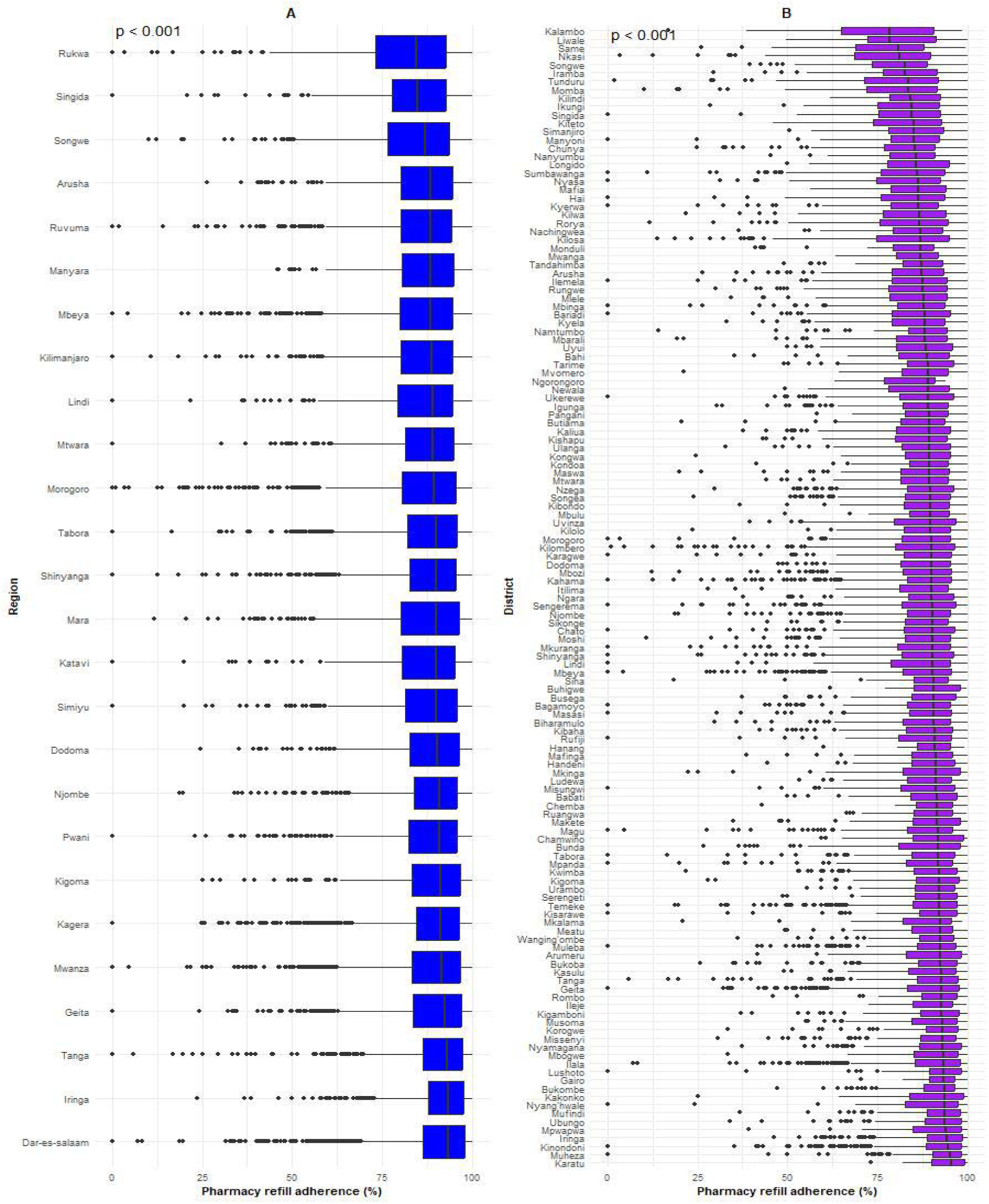
Boxplot of pharmacy refill adherence (%) among people living with HIV in Tanzania, 2017–2021: Panel A, regions; Panel B, districts, ordered by increasing median adherence.

### Association Between Adherence and Viral Suppression

Among the 21,572 patients with available viral load (VL) measurements, 88.7% achieved viral suppression (<1000 copies/mL). Suppression varied substantially across the demographic and clinical groups (Table 3). The rates were lowest among children (65.8%) and adolescents (74.4%), increased among young adults (89.9%), and were highest among adults aged 29–69 years (90.2%) and those aged ≥70 years (92.4%). Females had slightly higher suppression (89.2%) compared with males (87.8%). By marital status, suppression was highest among widowed (91.5%) and married patients (90.4%), while children had the lowest suppression (76.1%).

**Table 3:**
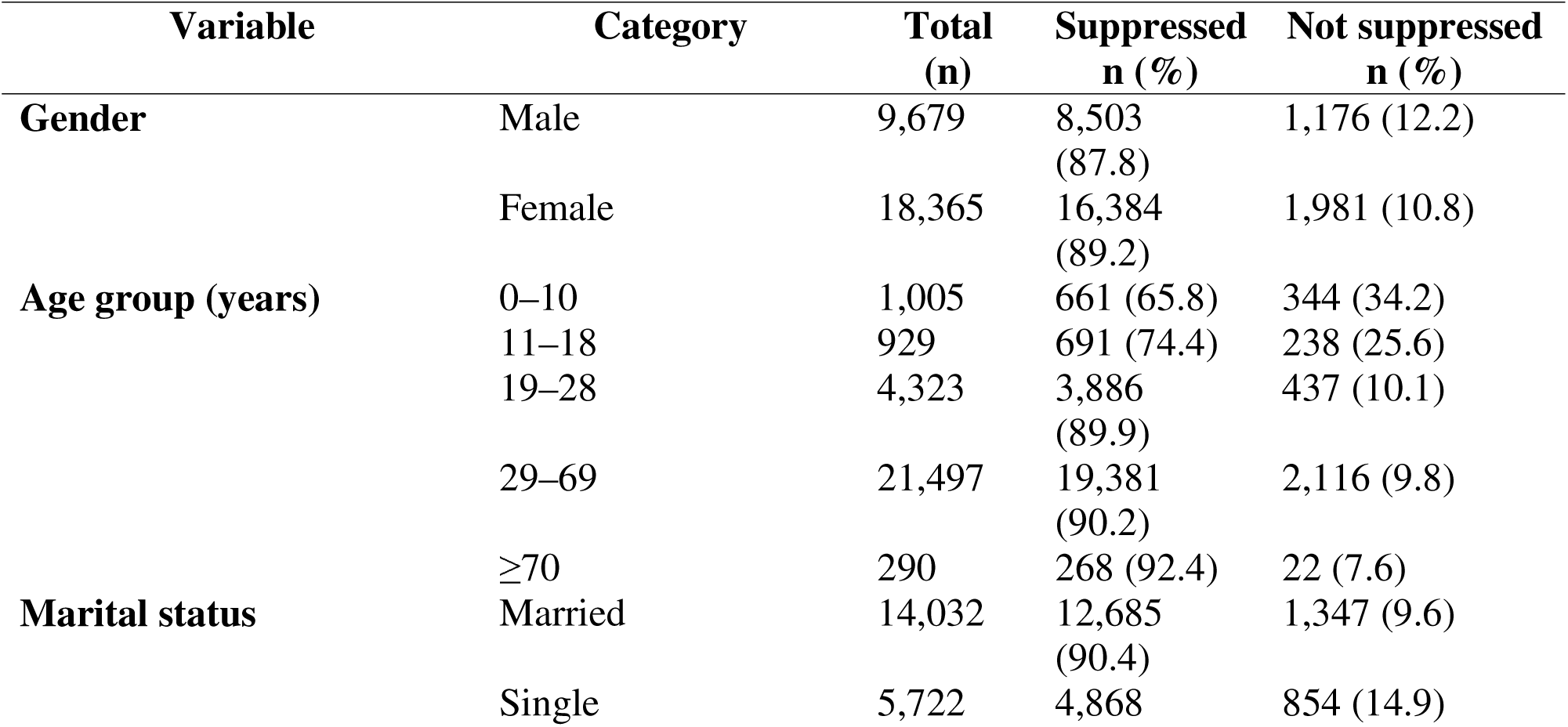
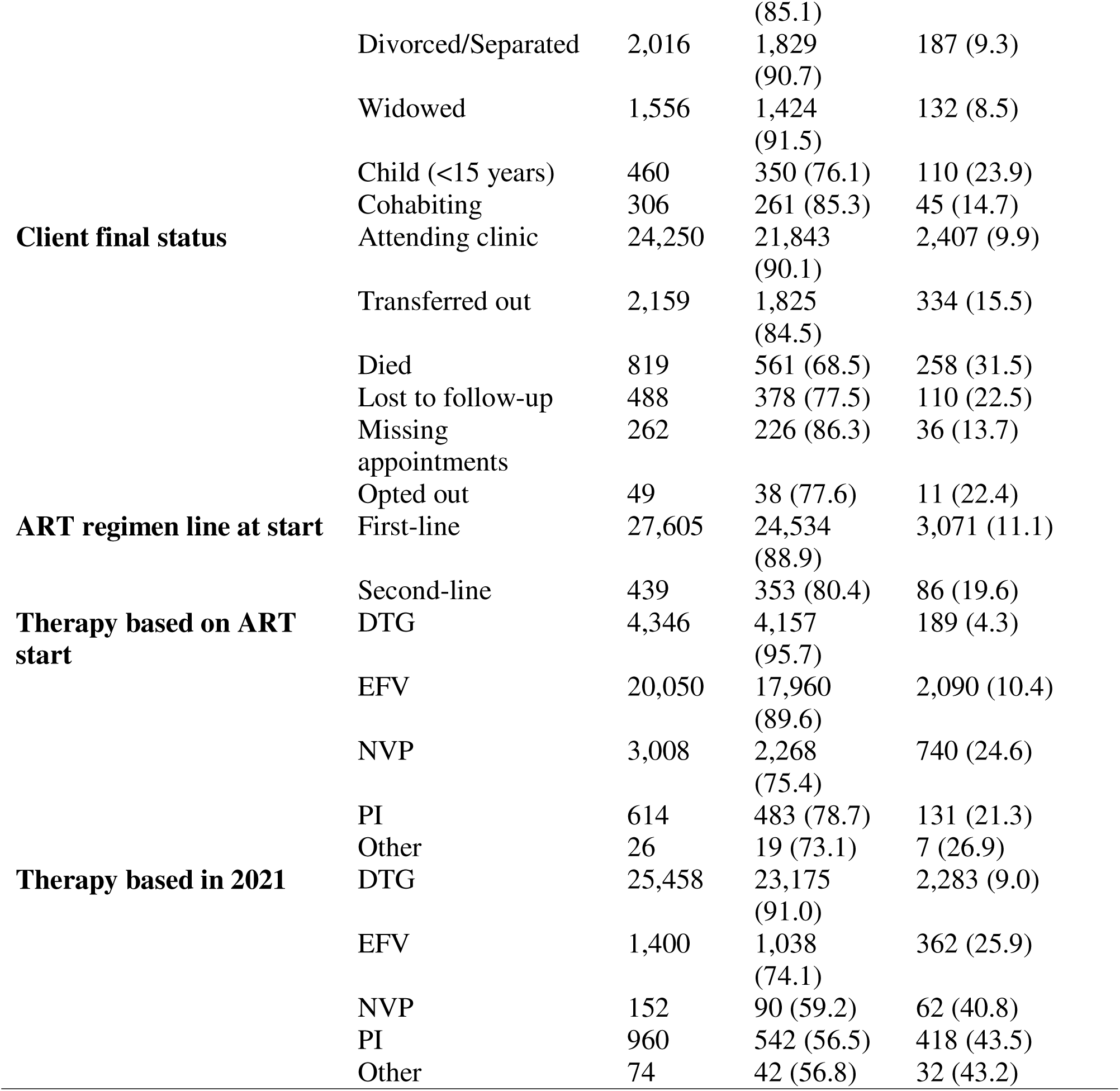
V**i**ral **load suppression (<1000 copies/mL) by demographic, clinical, and ART regimen characteristics**

Clinical engagement was strongly associated with the virological outcomes. Patients actively attending the same clinic had high suppression (90.1%), whereas lower suppression was observed among those transferred out (84.5%), lost to follow-up (77.5%), or reported as deceased (68.5%). Suppression also differed according to ART regimen characteristics. Patients initiated on first-line therapy had higher suppression (88.9%) than those on second-line therapy (80.4%). At ART initiation, DTG-based regimens showed the highest suppression (95.7%) compared with EFV-, NVP-, and PI-based regimens. By 2021, viral suppression remained highest among patients on DTG-based therapy (91.0%), while lower suppression was observed among those receiving NVP- or PI-based regimens.

Across all demographic, clinical, and regimen categories, higher pharmacy refill adherence was consistently associated with viral suppression, supporting its value as an objective indicator of treatment success (Table 3).

### .Machine Learning Model Performance

Pharmacy refill adherence consistently ranked among the strongest predictors of viral suppression across all evaluated models (Table 4), as measured by the area under the ROC curve and optimal adherence thresholds.

**Table 4:** Comparative model performance using the area under the receiver operating characteristic (ROC) curve (AUC) and optimal adherence thresholds for each approach.

| Model | AUC | Optimal adherence threshold (%) |
| --- | --- | --- |
| Logistic Regression | 0.62 | 88.2 |
| Random Forest | 0.60 | 98.8 |
| Gradient Boosting Machine | 0.66 | 89.3 |
| XGBoost | 0.67 | 88.3 |

XGBoost demonstrated the highest discriminative performance, followed by the Gradient Boosting Machine, whereas logistic regression provided a stable baseline, and Random Forest showed the lowest discrimination. Across the models, the optimal adherence thresholds clustered around 88–89%, except for the Random Forest model, which identified a higher threshold.

The ROC curves for the XGBoost models are shown in Figure 6. Panel A (without SMOTE) achieved an AUC of 0.676, whereas Panel B (with SMOTE) showed improved discrimination (AUC = 0.70), indicating enhanced detection of virological failure after class balancing.

**Figure 6:**
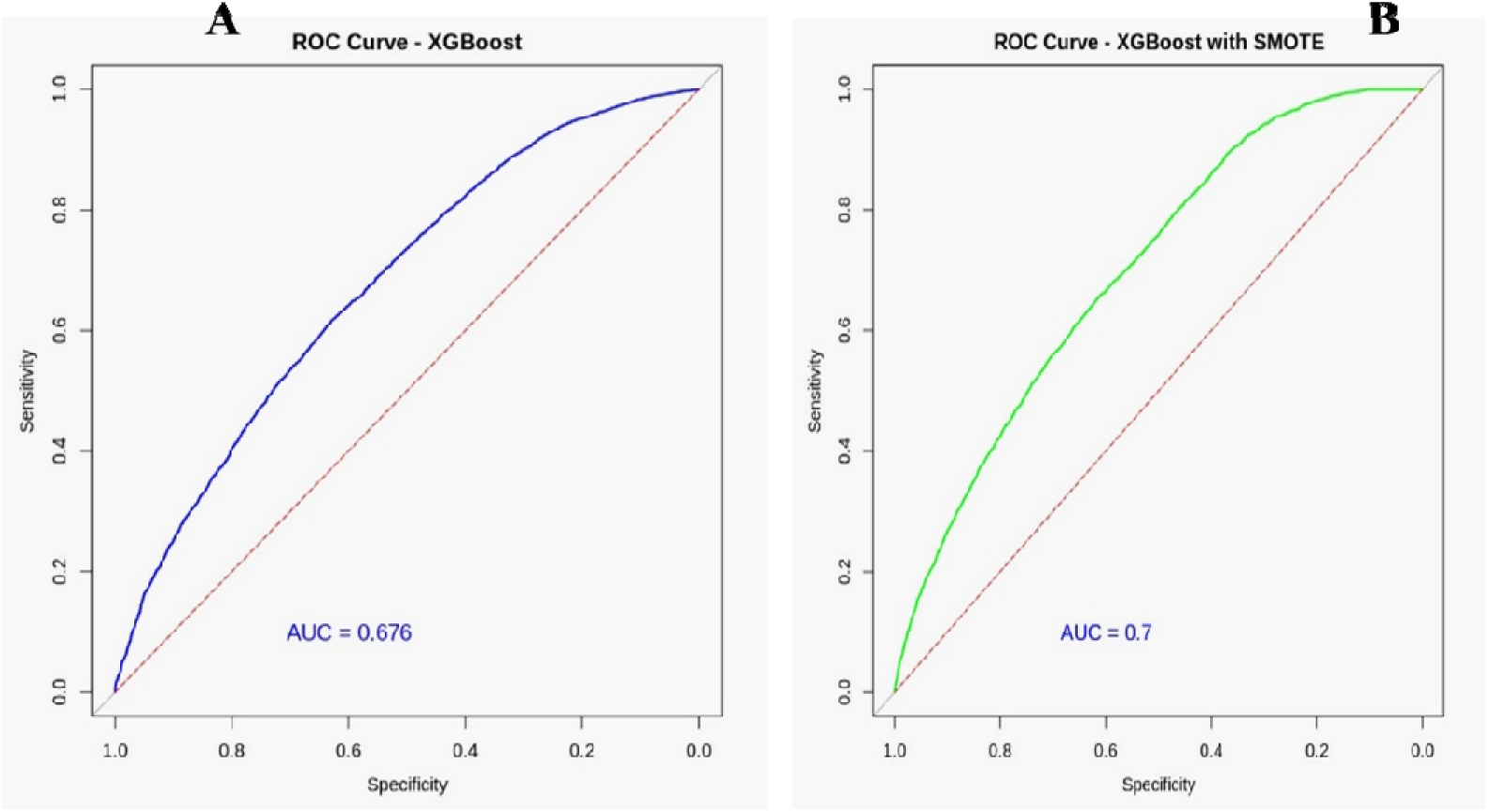
Receiver operating characteristic (ROC) curves (AUC) for XGBoost models: Panel A, without SMOTE; Panel B, with SMOTE.

### Factors Associated With Good Pharmacy Refill Adherence

Good pharmacy refill adherence (≥85%) varied according to demographic, clinical, and geographic characteristics (Table 5 and Figure 7). In the unadjusted analyses, lower adherence was observed among children, adolescents, and young adults than among adults aged 29–69 years. In contrast, higher adherence was observed among individuals with documented viral load (VL) measurements.

**Figure 7:**
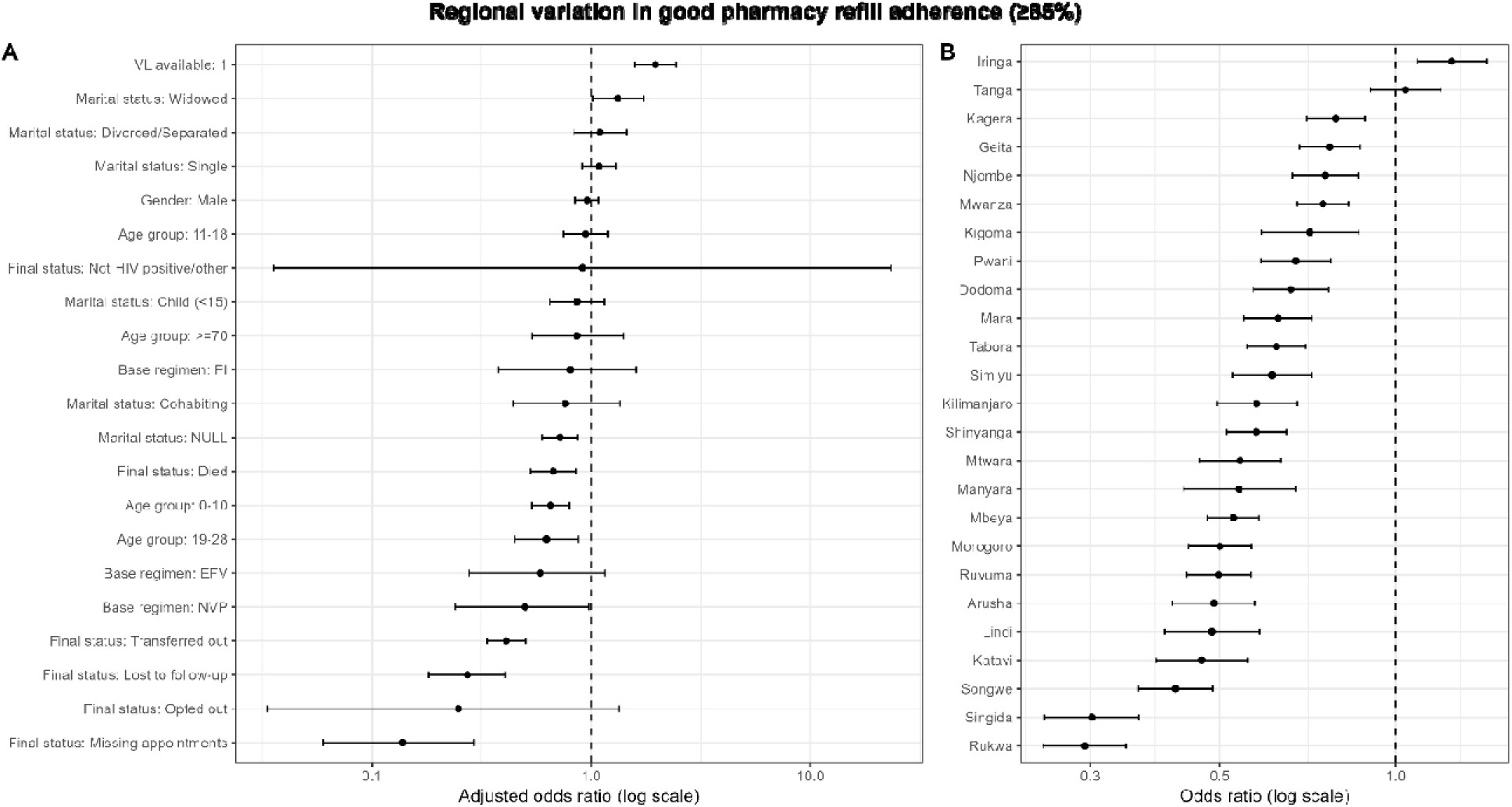
**Regional variation in good pharmacy refill adherence (**≥**85%).**

**Table 5:**
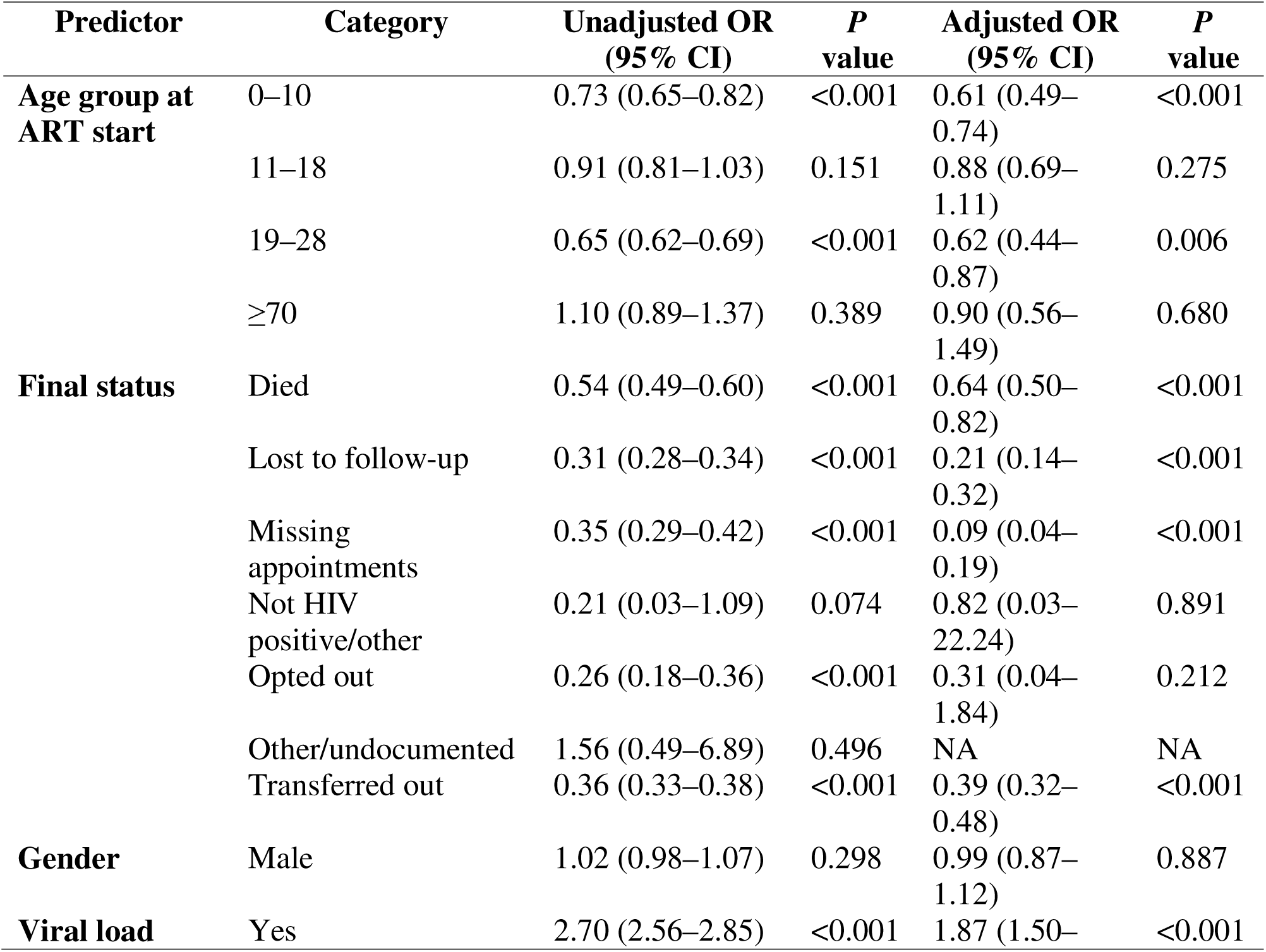
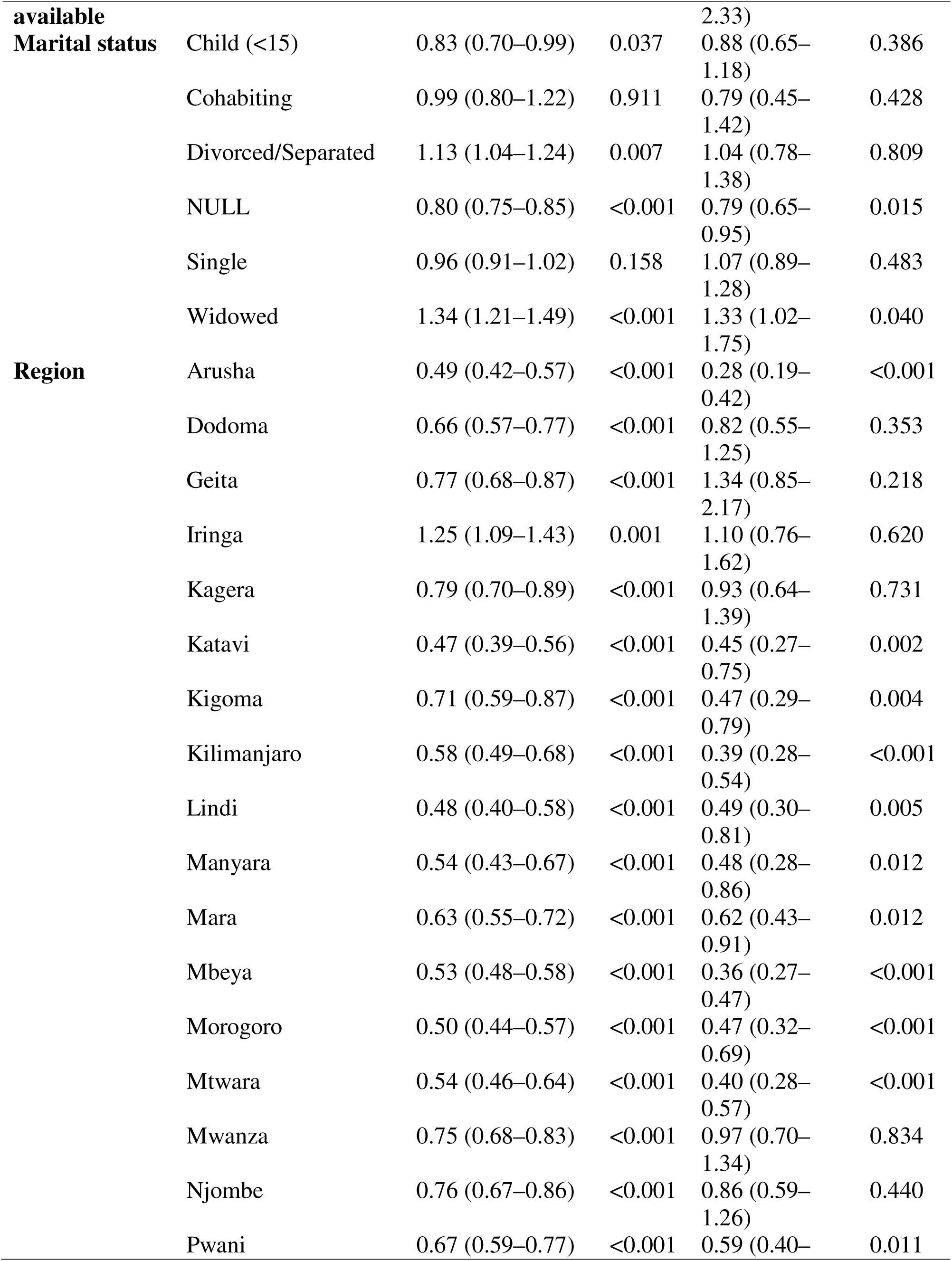
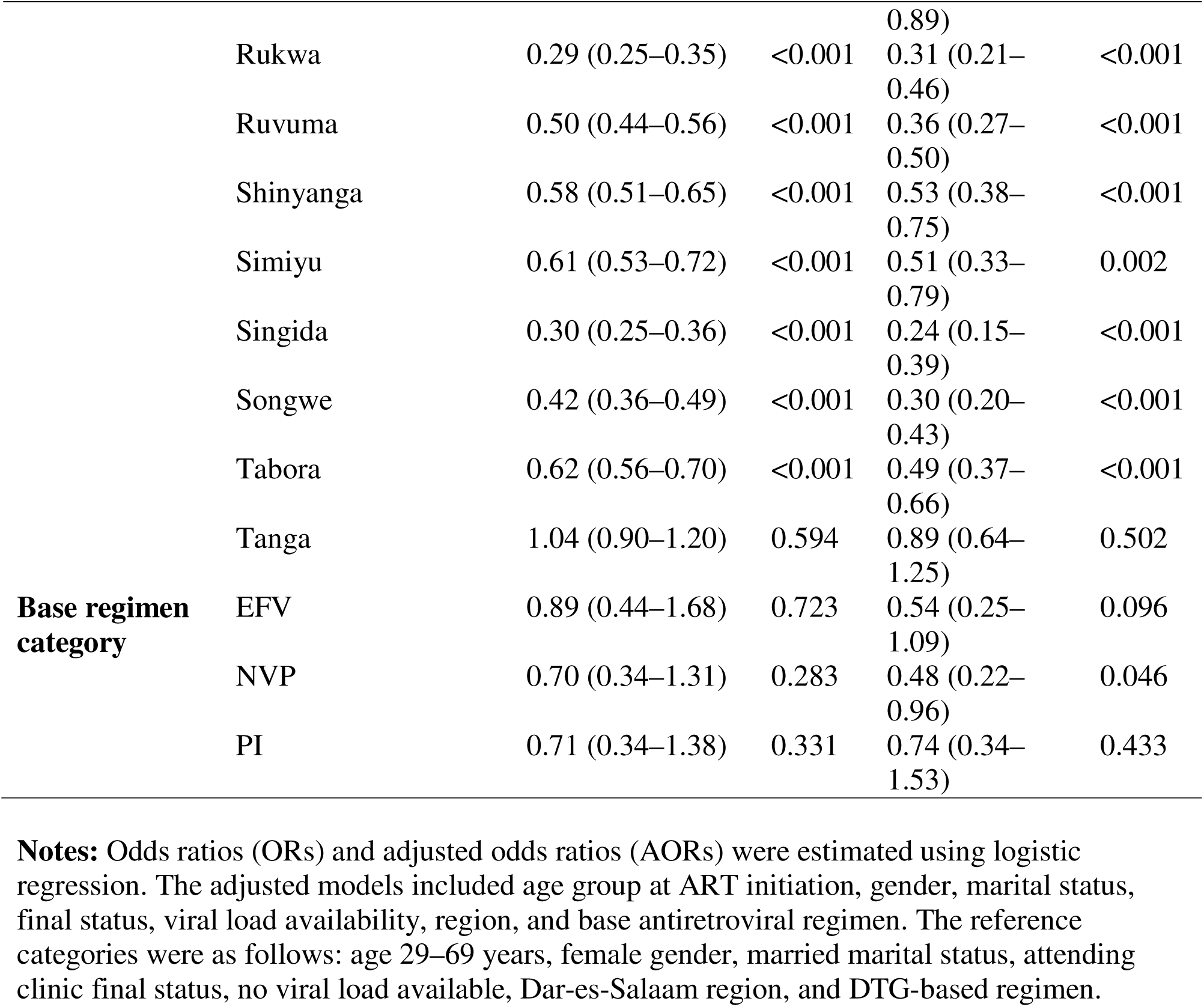
F**a**ctors **independently associated with good (**≥**85%) pharmacy refill adherence among people living with HIV (PLHIV).**

In the multivariable analysis, viral load availability remained independently associated with good adherence (adjusted odds ratio [AOR] 1.87; 95% CI 1.50–2.33). Compared with adults aged 29–69 years, children aged 0–10 years (AOR 0.61; 95% CI 0.49–0.74) and young adults aged 19–28 years (AOR 0.62; 95% CI 0.44–0.87) had significantly lower odds of good pharmacy refill adherence. Gender was not independently associated with adherence after adjustment.

The final patient status was strongly associated with adherence. Compared with patients actively attending clinic visits, those who were lost to follow-up, had missing appointments, or died had significantly lower adjusted odds of good adherence. Marital status was not consistently associated with adherence after adjustment, although widowed individuals had higher adjusted odds of adherence.

Substantial regional variation in adherence was observed (Figure 7). Compared with Dar es Salaam, multiple regions demonstrated significantly lower adjusted odds of good adherence, while no region showed significantly higher adjusted odds. Regarding antiretroviral regimens, patients on nevirapine-based regimens had lower adjusted odds of good adherence than those on dolutegravir-based regimens.

Panel A shows the adjusted odds ratios (AORs) for factors associated with good pharmacy refill adherence, excluding region, derived from a multivariable logistic regression model. Panel B shows the unadjusted odds ratios for regional variation in good pharmacy refill adherence, with Dar es Salaam as the reference category. The points represent odds ratios, and the horizontal lines indicate 95% confidence intervals. The vertical dashed line indicates an odds ratio of 1.0.

## Discussion

This national analysis demonstrates that pharmacy refill adherence, derived from routinely collected Care and Treatment Center (CTC) data, is strongly associated with viral suppression among people living with HIV (PLHIV) in Tanzania. Patients with higher refill adherence had a substantially greater likelihood of achieving virological suppression, whereas those with poor adherence were significantly more likely to experience virological failure. These findings are consistent with earlier studies from Tanzania and other resource-limited settings showing that objective pharmacy refill metrics outperform self-reported adherence measures in predicting treatment outcomes (Martin et al., 2017; Sangeda et al., 2014).

The overall viral suppression rate observed in this study aligns with national estimates and reflects progress toward achieving the UNAIDS 95–95–95 targets. However, persistent gaps in viral load (VL) testing, particularly among adolescents, young adults, and patients receiving care in remote facilities, highlight inequities that may undermine national progress. Similar disparities have been documented across sub-Saharan Africa, where logistical barriers, limited laboratory capacity, and inconsistent sample referral systems constrain VL coverage (Lecher et al., 2021; Roberts et al., 2016; Rutstein et al., 2016). In Tanzania, despite the adoption of routine VL monitoring policies and expansion of ART services (United Republic of Tanzania, 2019), structural and demographic barriers continue to influence access, as documented in prior studies from Tanzania and comparable settings (Lowenthal et al., 2014; Martelli et al., 2019)

A key contribution of this study is the demonstration that pharmacy refill adherence can function as a reliable proxy for treatment monitoring when VL testing is delayed or unavailable. The strong predictive performance of machine learning (ML) models, particularly gradient boosting and XGBoost, suggests that combining adherence metrics with other routinely collected programmatic variables, such as follow-up duration, visit frequency, and therapy change history, can enhance the early identification of patients at risk of treatment failure. This approach may be especially valuable in settings where laboratory-based monitoring remains inconsistent or resource-constrained. Prior modeling studies in southern Africa have emphasized the importance of supplementing VL testing with strategies to identify individuals at the highest risk of failure(Estill et al., 2018; Keiser et al., 2011). Our findings extend this evidence by demonstrating that Tanzania’s national CTC-2 data infrastructure is well-suited to support such predictive approaches at scale.

Several demographic patterns observed in this analysis warrant further attention. Adolescents and young adults exhibited consistently lower pharmacy refill adherence, in line with previous studies showing that adherence challenges in these groups are shaped by complex psychosocial factors, including caregiver dependence, stigma, disclosure difficulties, and increased mobility (Adejumo et al., 2015; Haberer & Mellins, 2009; Nichols et al., 2017). These vulnerabilities are associated with poorer viral suppression and an elevated risk of HIV drug resistance, as documented in Tanzanian and regional cohorts (Emmett et al., 2010; Muri et al., 2017; Tabb et al., 2018). The marked geographic variability in adherence and VL coverage observed across regions similarly reflects inequities reported in national surveys and programmatic assessments (United Republic of Tanzania, 2024; World Health Organization, 2025), underscoring the need for differentiated service delivery strategies tailored to local barriers.

The strong association between clinic engagement and pharmacy refill adherence underscores the central role of continuity of care in the management of chronic conditions, such as HIV. Patients who remained actively engaged in their care demonstrated substantially higher adherence, whereas those who were lost to follow-up or missed appointments exhibited the poorest refill adherence. These findings support the use of routine pharmacy refill monitoring as an early indicator of disengagement from care, allowing the timely identification of patients at risk of treatment interruption. Similar patterns have been reported across HIV programs in low-and middle-income countries, where pharmacy refill data are increasingly integrated into programmatic monitoring to trigger targeted adherence support and patient tracing interventions (Martin et al., 2017; Roberts et al., 2016).

This study also provides evidence supporting the feasibility of applying machine learning approaches to enhance decision-making within national HIV treatment programs. While logistic regression produced stable baseline estimates, ensemble methods, such as random forests and boosting algorithms, more effectively captured non-linear relationships and improved the classification of virological outcomes. As VL testing capacity continues to expand, integrating predictive analytics based on routine programmatic data could help optimize resource allocation, prioritize follow-up for patients at the greatest risk, and strengthen ART program performance.

Future work may explore sequence-based deep learning approaches, such as long short-term memory networks, to model longitudinal adherence trajectories using routine programmatic data (Hochreiter & Schmidhuber, 1997). In parallel, integrating these data with digital adherence tools informed by qualitative insights could enable scalable, people-centered strategies to improve ART adherence and viral suppression rates.

## Strengths and Limitations

A major strength of this study is the use of a large, nationally representative dataset covering diverse populations across 26 regions in Tanzania. The inclusion of pharmacy refill data collected over five years allowed a comprehensive assessment of adherence patterns and their association with virological outcomes. The use of multiple machine learning (ML) models further enabled the comparison of predictive performance using routinely collected programmatic data.

This study has several limitations. First, pharmacy refill adherence measures the collection of medication rather than actual ingestion. Although strongly associated with virological outcomes, it may overestimate adherence. Second, viral load (VL) testing was incomplete, which may have introduced bias if individuals without VL measurements systematically differed from those with available results. This limitation is common in analyses based on routine program data in resource-limited settings and highlights the importance of complementary monitoring approaches, such as refill-based adherence measures. Third, although ML models improved prediction performance, the analyses were restricted to variables available in routine databases; psychosocial, structural, and behavioral factors that may influence adherence and viral suppression were not included.

## Implications for Policy and Practice

These findings indicate that pharmacy refill adherence can serve as a cost-effective and scalable indicator of virological suppression in settings where VL testing coverage remains limited.

Incorporating ML-based adherence prediction into national HIV program monitoring systems could support the early identification of patients at an increased risk of treatment failure and enable more targeted adherence support. These approaches are consistent with Tanzania’s efforts to strengthen differentiated service delivery, expand digital health systems, and improve ART program performance in line with the UNAIDS 95–95–95 targets.

## Conclusion

Pharmacy refill adherence is a strong predictor of viral suppression among people living with HIV in Tanzania and is particularly valuable in contexts with delayed or incomplete VL testing.

When combined with machine learning models applied to routinely collected clinical and demographic data, refill adherence can support the early detection of treatment failure, inform targeted adherence interventions, and strengthen national HIV program monitoring. Scaling predictive analytic approaches within routine HIV care may contribute to improved long-term ART outcomes and support progress toward ending AIDS as a public health threat by 2030.

## Declarations Ethical Approval

Ethical clearance for this study was obtained from the Directorate of Research and Publication, Muhimbili University of Health and Allied Sciences (MUHAS), reference number **DA.282/298/01.C**. Permission to access and use the de-identified national CTC-2 database records was granted by the National AIDS, Sexually Transmitted Diseases, and Hepatitis Control Programme (NASHCOP). All methods were performed in accordance with the relevant national and international guidelines and regulations.

## Consent for Publication

Not applicable. This study used de-identified secondary data with no direct patient contact.

## Availability of Data and Materials

The Government of Tanzania owns the data used in this study through NASHCOP, and they are not publicly available. The de-identified analytical code and derived output can be made available from the corresponding author upon reasonable request and subject to NASHCOP approval.

## Authors’ Contributions

Meshack D. Lugoba, James Mwakyomo and Raphael Z. Sangeda conceived the study, designed the analytical framework, conducted data management and analysis, and interpreted the results. Ritah F. Mutagonda, George Musiba, Veryeh Sambu, Beatrice Mutayoba, Mercy Mpatwa, Prosper Njau, and Werner Maokola contributed to data acquisition, methodological input, and interpretation of findings. All authors participated in drafting and critically revising the manuscript for important intellectual content and approved the final version for publication.

## Acknowledgements

The authors acknowledge the Ministry of Health, NASHCOP leadership, and all healthcare workers across Tanzania’s Care and Treatment Centers for maintaining the national CTC-2 database.

## Supplementary Materials

**Supplementary Table 1.** Region Level Adherence (N = 26) in Tanzania

| Sno | Region | Median adherence |
| --- | --- | --- |
| 1 | Dar-es-salaam | 93.5 |
| 2 | Iringa | 93.4 |
| 3 | Tanga | 92.9 |
| 4 | Geita | 92.5 |
| 5 | Mwanza | 91.4 |
| 6 | Kagera | 91.3 |
| 7 | Kigoma | 91.2 |
| 8 | Pwani | 90.9 |
| 9 | Njombe | 90.8 |
| 10 | Dodoma | 90.4 |
| 11 | Simiyu | 90.1 |
| 12 | Katavi | 90.1 |
| 13 | Mara | 90.0 |
| 14 | Shinyanga | 90.0 |
| 15 | Tabora | 89.9 |
| 16 | Morogoro | 89.5 |
| 17 | Mtwara | 89.0 |
| 18 | Lindi | 89.0 |
| 19 | Kilimanjaro | 88.9 |
| 20 | Mbeya | 88.5 |
| 21 | Manyara | 88.5 |
| 22 | Ruvuma | 88.4 |
| 23 | Arusha | 88.3 |
| 24 | Songwe | 86.9 |
| 25 | Singida | 84.8 |
| 26 | Rukwa | 84.6 |

**Supplementary Table 2.**
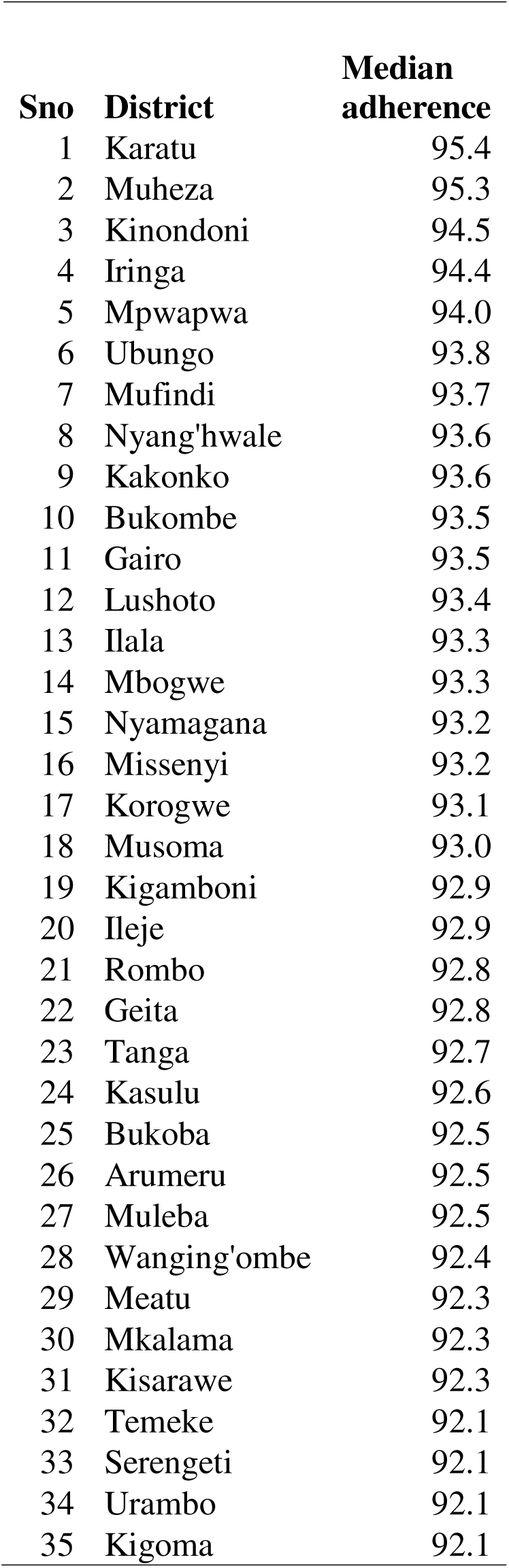
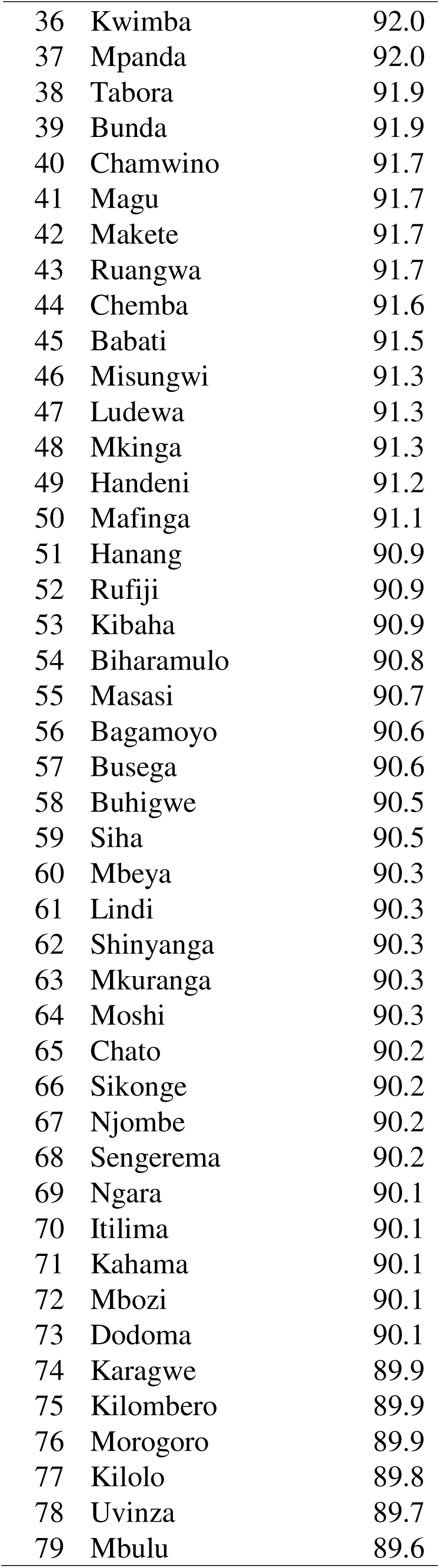
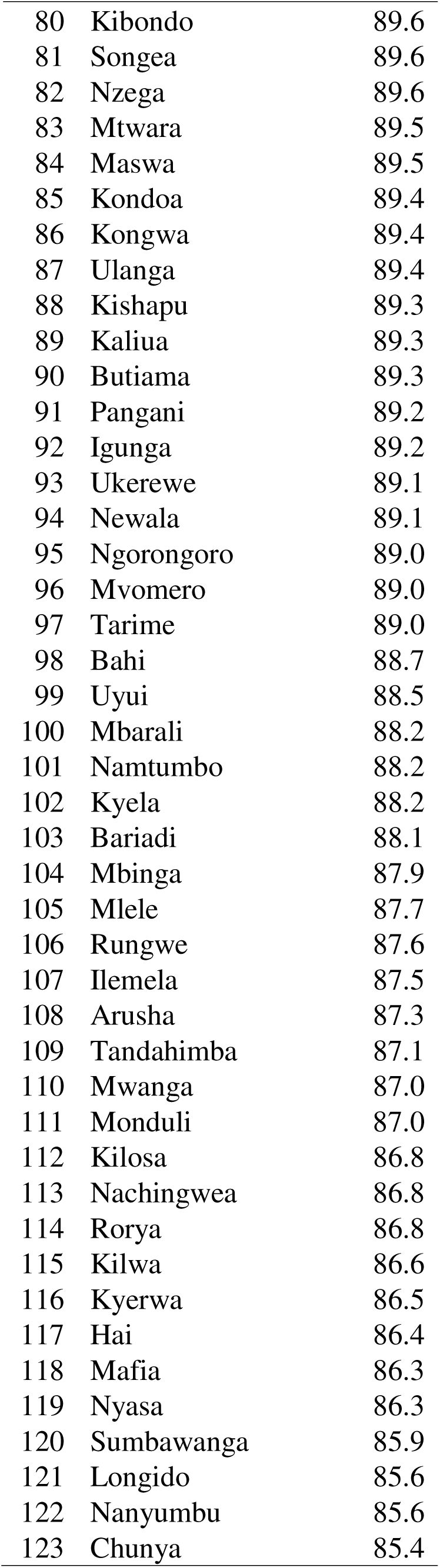
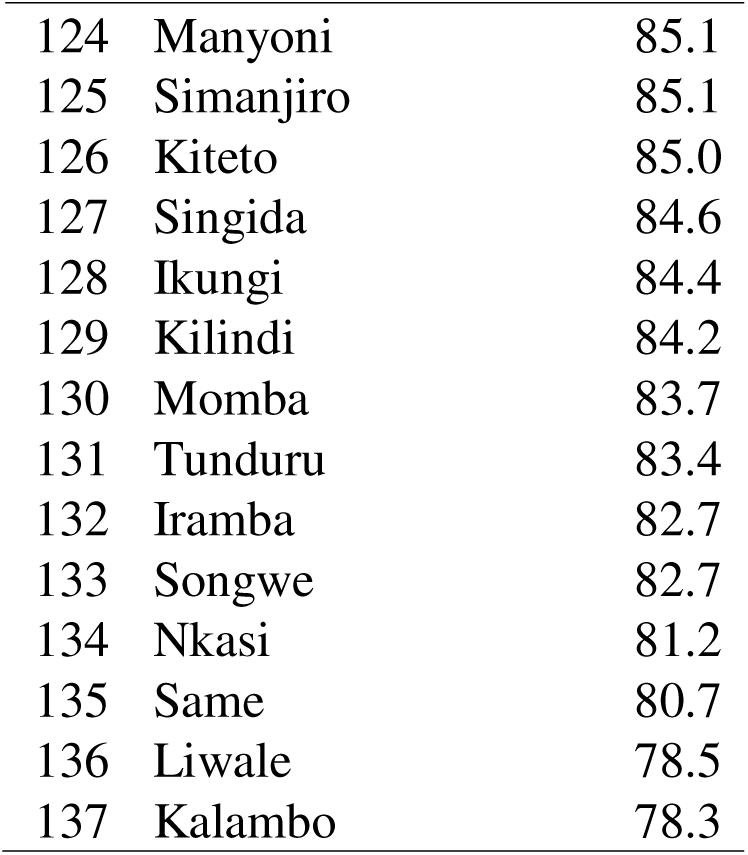
District Level Adherence (N = 137) in Tanzania

| **Sno** | **District** | **Median adherence** |
| --- | --- | --- |
| 1 | Karatu | 95.4 |
| 2 | Muheza | 95.3 |
| 3 | Kinondoni | 94.5 |
| 4 | Iringa | 94.4 |
| 5 | Mpwapwa | 94.0 |
| 6 | Ubungo | 93.8 |
| 7 | Mufindi | 93.7 |
| 8 | Nyang'hwale | 93.6 |
| 9 | Kakonko | 93.6 |
| 10 | Bukombe | 93.5 |
| 11 | Gairo | 93.5 |
| 12 | Lushoto | 93.4 |
| 13 | Ilala | 93.3 |
| 14 | Mbogwe | 93.3 |
| 15 | Nyamagana | 93.2 |
| 16 | Missenyi | 93.2 |
| 17 | Korogwe | 93.1 |
| 18 | Musoma | 93.0 |
| 19 | Kigamboni | 92.9 |
| 20 | Ileje | 92.9 |
| 21 | Rombo | 92.8 |
| 22 | Geita | 92.8 |
| 23 | Tanga | 92.7 |
| 24 | Kasulu | 92.6 |
| 25 | Bukoba | 92.5 |
| 26 | Arumeru | 92.5 |
| 27 | Muleba | 92.5 |
| 28 | Wanging'ombe | 92.4 |
| 29 | Meatu | 92.3 |
| 30 | Mkalama | 92.3 |
| 31 | Kisarawe | 92.3 |
| 32 | Temeke | 92.1 |
| 33 | Serengeti | 92.1 |
| 34 | Urambo | 92.1 |
| 35 | Kigoma | 92.1 |
| 36 | Kwimba | 92.0 |
| 37 | Mpanda | 92.0 |
| 38 | Tabora | 91.9 |
| 39 | Bunda | 91.9 |
| 40 | Chamwino | 91.7 |
| 41 | Magu | 91.7 |
| 42 | Makete | 91.7 |
| 43 | Ruangwa | 91.7 |
| 44 | Chemba | 91.6 |
| 45 | Babati | 91.5 |
| 46 | Misungwi | 91.3 |
| 47 | Ludewa | 91.3 |
| 48 | Mkinga | 91.3 |
| 49 | Handeni | 91.2 |
| 50 | Mafinga | 91.1 |
| 51 | Hanang | 90.9 |
| 52 | Rufiji | 90.9 |
| 53 | Kibaha | 90.9 |
| 54 | Biharamulo | 90.8 |
| 55 | Masasi | 90.7 |
| 56 | Bagamoyo | 90.6 |
| 57 | Busega | 90.6 |
| 58 | Buhigwe | 90.5 |
| 59 | Siha | 90.5 |
| 60 | Mbeya | 90.3 |
| 61 | Lindi | 90.3 |
| 62 | Shinyanga | 90.3 |
| 63 | Mkuranga | 90.3 |
| 64 | Moshi | 90.3 |
| 65 | Chato | 90.2 |
| 66 | Sikonge | 90.2 |
| 67 | Njombe | 90.2 |
| 68 | Sengerema | 90.2 |
| 69 | Ngara | 90.1 |
| 70 | Itilima | 90.1 |
| 71 | Kahama | 90.1 |
| 72 | Mbozi | 90.1 |
| 73 | Dodoma | 90.1 |
| 74 | Karagwe | 89.9 |
| 75 | Kilombero | 89.9 |
| 76 | Morogoro | 89.9 |
| 77 | Kilolo | 89.8 |
| 78 | Uvinza | 89.7 |
| 79 | Mbulu | 89.6 |
| 80 | Kibondo | 89.6 |
| 81 | Songea | 89.6 |
| 82 | Nzega | 89.6 |
| 83 | Mtwara | 89.5 |
| 84 | Maswa | 89.5 |
| 85 | Kondoa | 89.4 |
| 86 | Kongwa | 89.4 |
| 87 | Ulanga | 89.4 |
| 88 | Kishapu | 89.3 |
| 89 | Kaliua | 89.3 |
| 90 | Butiama | 89.3 |
| 91 | Pangani | 89.2 |
| 92 | Igunga | 89.2 |
| 93 | Ukerewe | 89.1 |
| 94 | Newala | 89.1 |
| 95 | Ngorongoro | 89.0 |
| 96 | Mvomero | 89.0 |
| 97 | Tarime | 89.0 |
| 98 | Bahi | 88.7 |
| 99 | Uyui | 88.5 |
| 100 | Mbarali | 88.2 |
| 101 | Namtumbo | 88.2 |
| 102 | Kyela | 88.2 |
| 103 | Bariadi | 88.1 |
| 104 | Mbinga | 87.9 |
| 105 | Mlele | 87.7 |
| 106 | Rungwe | 87.6 |
| 107 | Ilemela | 87.5 |
| 108 | Arusha | 87.3 |
| 109 | Tandahimba | 87.1 |
| 110 | Mwanga | 87.0 |
| 111 | Monduli | 87.0 |
| 112 | Kilosa | 86.8 |
| 113 | Nachingwea | 86.8 |
| 114 | Rorya | 86.8 |
| 115 | Kilwa | 86.6 |
| 116 | Kyerwa | 86.5 |
| 117 | Hai | 86.4 |
| 118 | Mafia | 86.3 |
| 119 | Nyasa | 86.3 |
| 120 | Sumbawanga | 85.9 |
| 121 | Longido | 85.6 |
| 122 | Nanyumbu | 85.6 |
| 123 | Chunya | 85.4 |
| 124 | Manyoni | 85.1 |
| 125 | Simanjiro | 85.1 |
| 126 | Kiteto | 85.0 |
| 127 | Singida | 84.6 |
| 128 | Ikungi | 84.4 |
| 129 | Kilindi | 84.2 |
| 130 | Momba | 83.7 |
| 131 | Tunduru | 83.4 |
| 132 | Iramba | 82.7 |
| 133 | Songwe | 82.7 |
| 134 | Nkasi | 81.2 |
| 135 | Same | 80.7 |
| 136 | Liwale | 78.5 |
| 137 | Kalambo | 78.3 |

## References

1. Adejumo, O. A., Malee, K. M., Ryscavage, P., Hunter, S. J., & Taiwo, B. O. (2015). Contemporary issues on the epidemiology and antiretroviral adherence of HIV-infected adolescents in sub-Saharan Africa: a narrative review. Journal of the International AIDS Society, 18(1), 20049. 10.7448/IAS.18.1.20049

2. Amour, M., Sangeda, R. Z., Kidenya, B., Balandya, E., Mmbaga, B. T., Machumi, L., Rugarabamu, A., Aris, E., Njiro, B. J., Ndumwa, H. P., Lyamuya, E., & Sunguya, B. F. (2022). Adherence to Antiretroviral Therapy by Medication Possession Ratio and Virological Suppression among Adolescents and Young Adults Living with HIV in Dar es Salaam, Tanzania. Tropical Medicine and Infectious Disease 2022, *Vol. 7, Page 52*, *7*(4), 52. 10.3390/TROPICALMED7040052

3. Breiman, L. (2001). Random Forests. Machine Learning, 45(1), 5–32. 10.1023/A:1010933404324

4. Chen, T., & Guestrin, C. (2016). XGBoost. *Proceedings of the 22nd ACM SIGKDD International Conference on Knowledge Discovery and Data Mining*, 785–794. 10.1145/2939672.2939785

5. Emmett, S. D., Cunningham, C. K., Mmbaga, B. T., Kinabo, G. D., Schimana, W., Swai, M. E., Bartlett, J. A., Crump, J. a, & Reddy, E. a. (2010). Predicting virologic failure among HIV-1-infected children receiving antiretroviral therapy in Tanzania: a cross-sectional study. Journal of Acquired Immune Deficiency Syndromes (1999), *54*(4), 368–375. 10.1097/QAI.0b013e3181cf4882

6. Estill, J., Kerr, C. C., Blaser, N., Salazar-Vizcaya, L., Tenthani, L., Wilson, D. P., & Keiser, O. (2018). The Effect of Monitoring Viral Load and Tracing Patients Lost to Follow-up on the Course of the HIV Epidemic in Malawi: A Mathematical Model. Open Forum Infectious Diseases, 5(5). 10.1093/ofid/ofy092

7. Haberer, J., & Mellins, C. (2009). Pediatric adherence to HIV antiretroviral therapy. Current HIV/AIDS Reports, 6(4), 194–200. 10.1007/s11904-009-0026-8

8. Hochreiter, S., & Schmidhuber, J. (1997). Long Short-Term Memory. Neural Computation, 9(8), 1735–1780. 10.1162/neco.1997.9.8.1735

9. Keiser, O., Chi, B. H., Gsponer, T., Boulle, A., Orrell, C., Phiri, S., Maxwell, N., Maskew, M., Prozesky, H., Fox, M. P., Westfall, A., & Egger, M. (2011). Outcomes of antiretroviral treatment in programmes with and without routine viral load monitoring in Southern Africa. AIDS (London, England), 25(14), 1761–1769. 10.1097/QAD.0b013e328349822f

10. Lecher, S. L., Fonjungo, P., Ellenberger, D., Toure, C. A., Alemnji, G., Bowen, N., Basiye, F., Beukes, A., Carmona, S., de Klerk, M., Diallo, K., Dziuban, E., Kiyaga, C., Mbah, H., Mengistu, J., Mots’oane, T., Mwangi, C., Mwangi, J. W., Mwasekaga, M., … Alexander, H. (2021). HIV Viral Load Monitoring Among Patients Receiving Antiretroviral Therapy — Eight Sub-Saharan Africa Countries, 2013–2018. MMWR. Morbidity and Mortality Weekly Report, 70(21), 775–778. 10.15585/mmwr.mm7021a2

11. Lowenthal, E. D., Bakeera-Kitaka, S., Marukutira, T., Chapman, J., Goldrath, K., & Ferrand, R. A. (2014). Perinatally acquired HIV infection in adolescents from sub-Saharan Africa: a review of emerging challenges. The Lancet Infectious Diseases, 14(7), 627–639. 10.1016/S1473-3099(13)70363-3

12. Mamo, D. N., Yilma, T. M., Tewelgne, M. F., Sebastian, Y., Bizuayehu, T., Melaku, M. S., & Walle, A. D. (2023). Machine learning to predict virological failure among HIV patients on antiretroviral therapy in the University of Gondar Comprehensive and Specialized Hospital, in Amhara Region, Ethiopia, 2022. BMC Medical Informatics and Decision Making, *23*(1), 75. 10.1186/s12911-023-02167-7

13. Martelli, G., Antonucci, R., Mukurasi, A., Zepherine, H., & Nöstlinger, C. (2019). Adherence to antiretroviral treatment among children and adolescents in Tanzania: Comparison between pill count and viral load outcomes in a rural context of Mwanza region. PLOS ONE, 14(3), e0214014. 10.1371/journal.pone.0214014

14. Martin, D., Luz, P. M., Lake, J. E., Clark, J. L., Campos, D. P., Veloso, V. G., Moreira, R. I., Cardoso, S. W., Klausner, J. D., & Grinsztejn, B. (2017). Pharmacy refill data can be used to predict virologic failure for patients on antiretroviral therapy in Brazil. Journal of the International AIDS Society, 20(1). 10.7448/IAS.20.1.21405

15. Muri, L., Gamell, A., Ntamatungiro, A. J., Glass, T. R., Luwanda, L. B., Battegay, M., Furrer, H., Hatz, C., Tanner, M., Felger, I., Klimkait, T., & Letang, E. (2017). Development of HIV drug resistance and therapeutic failure in children and adolescents in rural Tanzania. AIDS, 31(1), 61–70. 10.1097/QAD.0000000000001273

16. Nichols, J., Steinmetz, A., & Paintsil, E. (2017). Impact of HIV-Status Disclosure on Adherence to Antiretroviral Therapy Among HIV-Infected Children in Resource-Limited Settings: A Systematic Review. AIDS and Behavior, 21(1), 59–69. 10.1007/s10461-016-1481-z

17. Piot, P., & Quinn, T. C. (2013). Response to the AIDS Pandemic — A Global Health Model. New England Journal of Medicine, 368(23), 2210–2218. 10.1056/NEJMra1201533

18. Roberts, T., Cohn, J., Bonner, K., & Hargreaves, S. (2016). Scale-up of Routine Viral Load Testing in Resource-Poor Settings: Current and Future Implementation Challenges. Clinical Infectious Diseases, 62(8), 1043–1048. 10.1093/cid/ciw001

19. Rutstein, S. E., Golin, C. E., Wheeler, S. B., Kamwendo, D., Hosseinipour, M. C., Weinberger, M., Miller, W. C., Biddle, A. K., Soko, A., Mkandawire, M., Mwenda, R., Sarr, A., Gupta, S., & Mataya, R. (2016). On the front line of HIV virological monitoring: barriers and facilitators from a provider perspective in resource-limited settings. AIDS Care, 28(1), 1–10. 10.1080/09540121.2015.1058896

20. Sangeda, R. Z., Mosha, F., Prosperi, M., Aboud, S., Vercauteren, J., Camacho, R. J., Lyamuya, E. F., Van Wijngaerden, E., & Vandamme, A.-M. (2014). Pharmacy refill adherence outperforms self-reported methods in predicting HIV therapy outcome in resource-limited settings. BMC Public Health, 14(1), 1035. 10.1186/1471-2458-14-1035

21. Sollis, K. A., Smit, P. W., Fiscus, S., Ford, N., Vitoria, M., Essajee, S., Barnett, D., Cheng, B., Crowe, S. M., Denny, T., Landay, A., Stevens, W., Habiyambere, V., Perrins, J., & Peeling, R. W. (2014). Systematic Review of the Performance of HIV Viral Load Technologies on Plasma Samples. PLoS ONE, 9(2), e85869. 10.1371/journal.pone.0085869

22. Tabb, Z. J., Mmbaga, B. T., Gandhi, M., Louie, A., Kuncze, K., Okochi, H., Shayo, A. M., Turner, E. L., Cunningham, C. K., & Dow, D. E. (2018). Antiretroviral drug concentrations in hair are associated with virologic outcomes among young people living with HIV in Tanzania. AIDS, 32(9), 1115–1123. 10.1097/QAD.0000000000001788

23. UNAIDS. (2016). *Fast-track: ending the AIDS epidemic by* 2030. http://www.unaids.org/sites/default/files/media_asset/JC2686_WAD2014report_en.pdf

24. United Republic of Tanzania. (2019). National Guidelines for the management of HIV and AIDS in Tanzania. http://www.nacp.go.tz/download/national-guidelines-for-the-management-of-hiv-and-aids/

25. United Republic of Tanzania. (2024). *Tanzania HIV Impact Survey 2022-*2023. https://www.nbs.go.tz/nbs/takwimu/THIS2022-2023/THIS2022-2023_Summary_Sheet.pdf

26. World Health Organization. (2025). Tanzania HIV and AIDS. https://www.who.int/news-room/fact-sheets/detail/hiv-aids

27. Yang, X., Cai, R., Ma, Y., Zhang, H. H., Sun, X., Olatosi, B., Weissman, S., Li, X., & Zhang, J. (2025). Using Machine Learning Techniques to Predict Viral Suppression Among People With HIV. JAIDS Journal of Acquired Immune Deficiency Syndromes, 98(3), 209–216. 10.1097/QAI.0000000000003561

